# Social isolation and psychological distress among southern US college students in the era of COVID-19

**DOI:** 10.1101/2021.12.31.21268596

**Authors:** Danielle Giovenco, Bonnie E Shook-Sa, Bryant Hutson, Laurie Buchanan, Edwin B Fisher, Audrey Pettifor

**Author notes:** **Corresponding Author:** Danielle Giovenco, PhD.

## Abstract

**Objective:** To examine the prevalence of psychological distress and its association with social isolation among University of North Carolina Chapel Hill (UNC-CH) students.

**Methods:** A cross-sectional survey was emailed to all students in June 2020. Students reported self-isolating none, some, most, or all of the time and were screened for clinically significant symptoms of depression (CSSD). Data were weighted to the UNC-CH population.

**Results:** 7,012 students completed surveys-64% reported self-isolating most or all of the time and 64% reported CSSD. Compared to those self-isolating none of the time, students self-isolating some of the time were 1.78 (95% CI 1.37-2.30) times as likely to report CSSD, and students self-isolating most and all of the time were 2.12 (95% CI 1.64-2.74) and 2.27 (95% CI 1.75-2.94) times as likely to report CSSD, respectively.

**Conclusions:** Universities should prioritize student mental health and prepare support services to mitigate mental health consequences of the pandemic.

## INTRODUCTION

The COVID-19 pandemic has resulted in poor mental health globally. In June 2020, a survey assessing mental health challenges related to COVID-19 among a nationally-representative sample of adults in the United States found that young adults (18-24 years) reported the highest prevalence of symptoms of anxiety (49%) and depression (52%), in addition to substance use to cope with pandemic-related stress (25%) and suicidal ideation (26%), compared to any other age group (1). The prevalence of adverse mental health outcomes among those aged 25-44-years was also high, with prevalence decreasing as age increased. College students are a unique group of young adults facing academic, interpersonal, and environmental stressors, who have historically experienced high rates of mental health distress compared to the general population (2–4).

To limit the spread of SARS-CoV-2, colleges across the United States abruptly transitioned to remote learning in March 2020, closing their campuses and forcing students to learn remotely. During periods of social isolation and uncertainty, individuals are prone to experiencing heightened levels of psychological distress that have the potential to exacerbate a co-epidemic of mental health disorders and COVID-19 (5). While many students have since resumed in-person learning, prevention and control measures, such as physical distancing, will continue into 2022 (6). Thus, it is important to understand the impacts of social isolation on student mental health burden to inform university support systems. In the present study, we aimed to: 1) characterize the prevalence of symptoms of psychological distress among a large sample of public university students in the southern United States and 2) examine social isolation as a potential driver of psychological distress.

## METHODS

### Study overview

The University of North Carolina at Chapel Hill (UNC-CH) is a large public research university with 21,223 undergraduate and 12,016 graduate and professional degree-seeking students enrolled at the time of this study. In an initial response to COVID-19, UNC-CH significantly reduced operations on March 20, 2020, requiring students to vacate campus housing by March 21, and shifted to remote instruction on March 23.

A cross-sectional survey aimed at assessing student knowledge, attitudes, and behaviors related to SARS CoV-2 was emailed to all UNC-CH undergraduate, graduate, and professional students on June 8, 2020. The survey was open for two weeks until June 23. Prior to starting the survey, students interested in participating followed a link to read and sign an informed consent. The survey consisted of 47 questions, many of which had several parts, and incorporated multiple choice, multiple answer, and open-ended response questions. Questions related to COVID-19 were drawn from similar surveys or were based on our own design. We also included several validated measures to assess student well-being. The survey took approximately 30 minutes to complete. Students who completed at least 75% of the survey were entered into a drawing for one of fifty, $50 gift certificates.

Demographic and student data (e.g., graduate or undergraduate student type, full or part-time status, etc.) was provided by the university registrar and linked to survey responses prior to data de-identification for analyses. The Institutional Review Board of the UNC-CH Office of Human Research Ethics approved study procedures.

### Measures

#### Social Isolation exposures

For our primary exposure, participants were asked, “to what extent are you self-isolating?”. Answer options included: all of the time – I am staying at home nearly all the time; most of the time – I only leave my home to buy food and other essentials; some of the time – I have reduced the amount of times I am in public spaces, social gatherings, or at work; and none of the time – I am doing everything I normally do.

Several additional questions were used to assess social isolation. In a measure aimed at assessing attitudes towards COVID-19 prevention and control measures, participants were asked, “How much you disagree or agree with the following statements: 1) I avoid crowded areas and 2) I avoid getting together with people who are not part of my household”. In a measure aimed at assessing behavioral changes related to COVID-19, participants were asked, “To what extent do you agree with each of the following statements about your behavior in the past month as a result of the new coronavirus: 1) I stayed at home and 2) I did not attend social gatherings”. For both measures, participants were asked to rate their behavior on a 5-point Likert scale ranging from strongly disagree to strongly agree.

#### Psychological distress outcomes

Scores for each psychological distress outcome variable were calculated only for participants with complete data for all measure items.

The 10-item Center for Epidemiological Studies Depression Scale (CES-D-10) is a widely used questionnaire assessing clinically significant depressive symptoms in the previous week (7). It includes three items on depressed affect, five items on somatic symptoms, and two on positive affect. Likert scale options for each item range from “rarely or none of the time” (score of 0) to “all of the time” (score of 3). Scoring is reversed for items based on statements of positive affect. The total score is the sum of 10 items (possible range = 0-30). Based on previous studies (7), a total score equal to or above 10 was used to identify individuals reporting clinically significant symptoms of depression.

The 3-item Loneliness Scale (UCLA-3) is a questionnaire developed from the Revised UCLA Loneliness Scale assessing feelings of loneliness or social isolation in the previous month (8). Each question was rated on a 3-point scale: 1 = hardly ever; 2 = some of the time; 3 = often. All items are summed to give a total score with higher scores indicating greater degrees of loneliness (possible range = 3-9). Consistent with previous research, we categorized individuals with total scores equal to or above 6 as lonely (9,10).

The 4-item Perceived Stress Scale (PSS-4) is a questionnaire that assesses the degree to which situations in one’s life over the previous month are appraised as stressful (11,12). Each question was rated on a 5-point scale: 0 = never, 1 = almost never, 2 =sometimes, 3 = fairly often, 4 = very often. Scores are obtained by reverse coding two positive items and then summing scores across all 4 items with higher scores indicating a higher perceived stress level (possible range = 0-16). For analysis, total scores were dichotomized at the unweighted sample median, with total scores at or below the median indicating lower perceived stress and scores above the median indicating greater perceived stress.

### Analysis

Analyses included students who completed the survey, regardless of whether items were skipped. To adjust for student nonresponse, we used iterative proportional fitting (i.e., raking) methods to weight the sample of survey respondents to the marginal distributions of the UNC-CH student population by age category, race and ethnicity, gender, and student type (undergraduate or graduate/professional). University registrar data for all eligible students enrolled in June 2020 were used to create marginal control totals that were entered into the raking algorithm (13). Iterative weight adjustments continued until the weighted margins differed from population margins by <1% for each raking variable.

We described the unweighted and weighted sample distributions for demographic and student characteristics provided by the UNC-CH registrar. All following results were presented weighted, with their unweighted counterparts included in the appendix. First, the proportion of students who self-reported that they were self-isolating most or all of the time (vs. some or none of the time) was described for each level of demographic and student characteristics, and Wald chi-square tests were used to identify significant differences. Then, the overall prevalence of each social isolation exposure variable was described. Next, we described the overall prevalence of clinically significant depressive symptoms and loneliness, and the distribution of perceived stress. Wald chi-square tests were used to determine if there were significant differences in the prevalence of psychological distress outcomes by age, race/ethnicity, gender, and student type. We assessed the internal reliability of each outcome measure (CES-D-10, UCLA-3, PSS-4) using Cronbach’s alpha.

Log-binomial regression was used to calculate prevalence ratio (PR) estimates for associations between social isolation and psychological distress. Robust variance estimators were used for weighted regression models. For our primary exposure, we estimated the relative prevalence of each psychological distress outcome among participants who reported self-isolating some, most, or all of the time versus none of the time (referent), and a test for linear trend was conducted (α=.05). For each additional social isolation exposure, psychological distress prevalence among participants who selected “somewhat agree” or “strongly agree” was compared to participants who selected “somewhat disagree” or “strongly disagree” (referent). Statistical analyses were conducted in SAS 9.4 (Cary, NC).

## RESULTS

A total of 33,239 UNC-CH students were emailed the survey. Among these, 9,531 students started the survey (29% response rate), of whom 7,012 (74%) completed the survey and were included in this analysis. The distribution of respondents was largely similar to the distribution of UNC-CH students for demographic and student characteristic domains examined, with the exception of gender (Table 1). Thus, differences between weighted and unweighted estimates were minimal. The weighted student population was predominately <25 years of age (73%), female (58%), non-Hispanic white (61%), and enrolled in full-time study (69%). Sixty-four percent self-reported they were self-isolating most or all of the time.

**Table 1.** Demographic and student characteristics

| Characteristic | Unweighted sample | Weighted sample | Self-isolating most or all of the time<br>(versus some or none of the time) |  |  |
| --- | --- | --- | --- | --- | --- |
|  | N (%) | N (%) | Weighted N (%) | 95% CI | p-value<sup>c</sup> |
| **Total N** | 7012 | 33239 | 21251 (64%) | 62.8%, 65.2% |  |
| **Age** |  |  |  |  |  |
| <21 years | 3656 (52%) | 15497 (47%) | 9048 (58%) | 56.8%, 60.1% |  |
| 21-24 years | 1712 (24%) | 8548 (26%) | 5437 (64%) | 61.4%, 66.1% |  |
| 25-34 years | 1338 (19%) | 7066 (21%) | 5171 (73%) | 70.7%, 75.7% |  |
| ≥35 years | 305 (4%) | 2127 (6%) | 1595 (75%) | 70.0%, 80.3% | <.001 |
| **Race/Ethnicity** |  |  |  |  |  |
| White | 4422 (66%) | 19304 (61%) | 11078 (57%) | 56.0%, 59.0% |  |
| Black or African American | 406 (6%) | 2639 (8%) | 1921 (73%) | 68.1%, 77.4% |  |
| Asian | 1016 (15%) | 5323 (17%) | 4215 (79%) | 76.6%, 81.8% |  |
| Hispanic of any race | 541 (8%) | 2885 (9%) | 1972 (68%) | 64.4%, 72.5% |  |
| Other<sup>a</sup> or multiple races | 365 (5%) | 1724 (5%) | 1117 (65%) | 59.7%, 69.9% | <.001 |
| **Gender** |  |  |  |  |  |
| Female | 4999 (71%) | 19425 (58%) | 12602 (65%) | 63.6%, 66.3% |  |
| Male | 2007 (29%) | 13789 (42%) | 8628 (63%) | 60.5%, 64.8% | .080 |
| **Academic career** |  |  |  |  |  |
| Undergraduate student<sup>b</sup> | 4754 (68%) | 21223 (64%) | 12780 (60%) | 58.8%, 61.8% |  |
| Graduate/prof student | 2258 (32%) | 12016 (36%) | 8471 (71%) | 68.6%, 72.5% | <.001 |
| **Full-time status** |  |  |  |  |  |
| Part-time | 2206 (31%) | 10359 (31%) | 6325 (61%) | 58.9%, 63.2% |  |
| Full-time | 4805 (69%) | 22879 (69%) | 14925 (65%) | 63.9%, 66.8% | .001 |
| **Residency** |  |  |  |  |  |
| In-state | 5237 (75%) | 24176 (73%) | 15187 (63%) | 61.6%, 64.3% |  |
| Out-of-state | 1768 (25%) | 9028 (27%) | 6044 (67%) | 64.6%, 69.3% | .003 |
| **Citizenship** |  |  |  |  |  |
| U.S. citizen | 6376 (91%) | 29714 (90%) | 18448 (62%) | 60.9%, 63.4% |  |
| Non-U.S. citizen | 628 (9%) | 3485 (10%) | 2771 (80%) | 76.2%, 82.9% | <.001 |

Estimates exclude 1 (.01%) participant missing age, 262 (3.7%) missing race/ethnicity, 6 (.09%) missing gender, 1 (.01%) missing full-time status, 7 (.10%) missing residency, and 8 (.11%) missing citizenship.
<sup>a</sup>Includes 'American Indian or Alaskan Native' or 'Native Hawaiian or other Pacific Islander'
<sup>b</sup>Includes 18 post-baccalaureate students
<sup>c</sup>Wald chi-square test comparing self-isolation most or all of the time to some or none of the time

Self-isolation varied by demographic and student characteristics. Students 25-34 (73%) and ≥35 (75%) years were more likely to report they were self-isolating most or all of the time than those <21 (58%) and 21-24 (64%) years. Further, Asian and Black/African American students were most likely to be self-isolating (79% and 73%, respectively) than any other race group, and White race students were least likely (57%). Those who reported self-isolating were also more likely to be graduate/professional students, full-time students, out-of-state residents, and non-US citizens (Table 1). The majority of students agreed or strongly agreed with statements that they were avoiding crowded areas (97%) and not getting together with people outside of their households (79%), and that in the previous month they stayed home (93%) and did not attend social gatherings (90%) (Table 2).

**Table 2.** Weighted distribution of social isolation exposure variables (N=33,239)

| **Social isolation variables** | **N (%)** | **95% CI** |
| --- | --- | --- |
| **Self-isolation<sup>a</sup>** |  |  |
| None of the time | 659 (2%) | 1.6%, 2.3% |
| Some of the time | 11288 (34%) | 32.8%, 35.2% |
| Most of the time | 16927 (51%) | 49.8%, 52.2% |
| All of the time | 4324 (13%) | 12.2%, 13.9% |
| **I avoid crowded areas<sup>b</sup>** |  |  |
| Somewhat/strongly disagree | 1065 (3%) | 2.9%, 3.8% |
| Somewhat/strongly agree | 30890 (97%) | 96.2%, 97.1% |
| **I avoid getting together with people who are not part of my household<sup>b</sup>** |  |  |
| Somewhat/strongly disagree | 6416 (21%) | 20.4%, 22.5% |
| Somewhat/strongly agree | 23459 (79%) | 77.5%, 79.6% |
| **I stayed home<sup>c</sup>** |  |  |
| Somewhat/strongly disagree | 2352 (7%) | 6.6%, 7.9% |
| Somewhat/strongly agree | 29931 (93%) | 92.1%, 93.4% |
| **I did not attend social gatherings<sup>c</sup>** |  |  |
| Somewhat/strongly disagree | 3046 (10%) | 8.8%, 10.3% |
| Somewhat/strongly agree | 28724 (90%) | 89.7%, 91.2% |

Estimates exclude 8 (.11%) participants missing self-isolation; 21 (.30%) missing and 245 (3.5%) who responded "neither agree nor disagree" for "I avoid crowded areas"; 16 (.23%) missing and 705 (10.1%) who responded "neither agree nor disagree" for "I avoid getting together with people who are not in my household"; 14 (.20%) missing and 187 (2.7%) who responded "neither agree nor disagree" for "I stayed home"; and 25 (.36%) missing and 287 (4.1%) who responded "neither agree nor disagree" for "I did not attend social gatherings"
<sup>a</sup>To what extent are you self-isolating?
<sup>b</sup>Please indicate how much you disagree or agree with the following statements
<sup>c</sup>To what extent do you agree with each of the following statements about your behavior in the past month as a result of the new coronavirus?

Almost two-thirds (64%) of UNC-CH students reported clinically significant depressive symptoms and 65% were categorized as lonely (Table 3). Further, 41% of students reported perceived stress scores above the unweighted sample median for the PSS-4 scale of 8, indicating greater perceived stress. For the weighted sample, the median CES-D-10 score was 12 (IQR=7-17), the median UCLA-3 score was 6 (IQR=5-8), and the median PSS-4 score was 8 (IQR=6-10). All three psychological distress scales had good or acceptable internal consistency (CES-D-10 α=.87, UCLA-3 α=.78, and PSS-4 α=.76).

**Table 3.** Weighted distribution of psychological distress outcome variables (N=33,239)

| **Mental health variables** | **N (%)** | **95% CI** |
| --- | --- | --- |
| **CES-D-10** |  |  |
| Non-clinically significant depressive symptoms (score <10) | 11920 (36%) | 35.3%, 37.7% |
| Clinically significant depressive symptoms (score ≥10) | 20759 (64%) | 62.3%, 64.7% |
| **UCLA-3** |  |  |
| Not lonely (score=3-5) | 11702 (35%) | 34.1%, 36.5% |
| Lonely (score=6-9) | 21403 (65%) | 63.5%, 65.9% |
| **PSS-4** |  |  |
| Lower stress (score=0-8) | 19389 (59%) | 57.5%, 60.0% |
| Greater stress (score=9-16) | 13610 (41%) | 40.0%, 42.5% |

Estimates exclude 114 (1.6%) participants missing a CES-D-10 score, 24 (.34%) missing a UCLA-3 score, and 44 (.63%) missing a PSS-4 score; scores for the Perceived Stress Scale (PSS-4) were dichotomized at the median

Missing data were minimal, with <2% of students missing data for measure items.

Psychological distress outcome prevalence varied by age, race, gender, and student type. Women were more likely than men to report clinically significant depressive symptoms (71% vs. 54%, p<.001), loneliness (67% vs. 61%, p<.001), and greater perceived stress (48% vs. 31%, p<.001). Black/African American, Hispanic, and other/multiple race students were more likely than White and Asian students to report clinically significant depressive symptoms (66% vs. 63%, p=.032) and greater perceived stress (44% vs. 40%, p=.008). Clinically significant depressive symptoms varied significantly by age group (p=.011), with students 21-24 years reporting the highest prevalence (67%) and lower estimates in the other age groups (<21 years=62%, 25-34 years=63%, and ≥35 years=59%). Further, students <21 and 21-24 years were more likely than students 25-34 and ≥35 years to report loneliness (70% vs. 51%, p<.001) and greater perceived stress (43% vs. 36%, p<.001). Lastly, undergraduates were more likely than graduate/professional students to report loneliness (70% vs. 55%, p<.001) and greater perceived stress (43% vs. 37%, p<.001).

Self-isolation was significantly associated with prevalence of clinically significant depressive symptoms, loneliness, and greater perceived stress, such that a higher relative prevalence was observed for each increase in level of self-isolation (Figure 1). For example, compared to students self-isolating none of the time, students self-isolating some of the time were 1.78 times as likely to have clinically significant depressive symptoms (95% CI 1.37, 2.30). Further, students self-isolating most or all of the time were 2.12 (95% CI 1.64, 2.74) and 2.27 (95% CI 1.75, 2.94) times as likely to have clinically significant depressive symptoms, respectively. Significant linear trends (p<.001) were observed between level of social isolation and both clinically significant depressive symptoms and greater perceived stress. Further, we found similar associations between agreement with additional self-isolation statements and psychological distress outcome prevalence (Table 4).

**Table 4.**
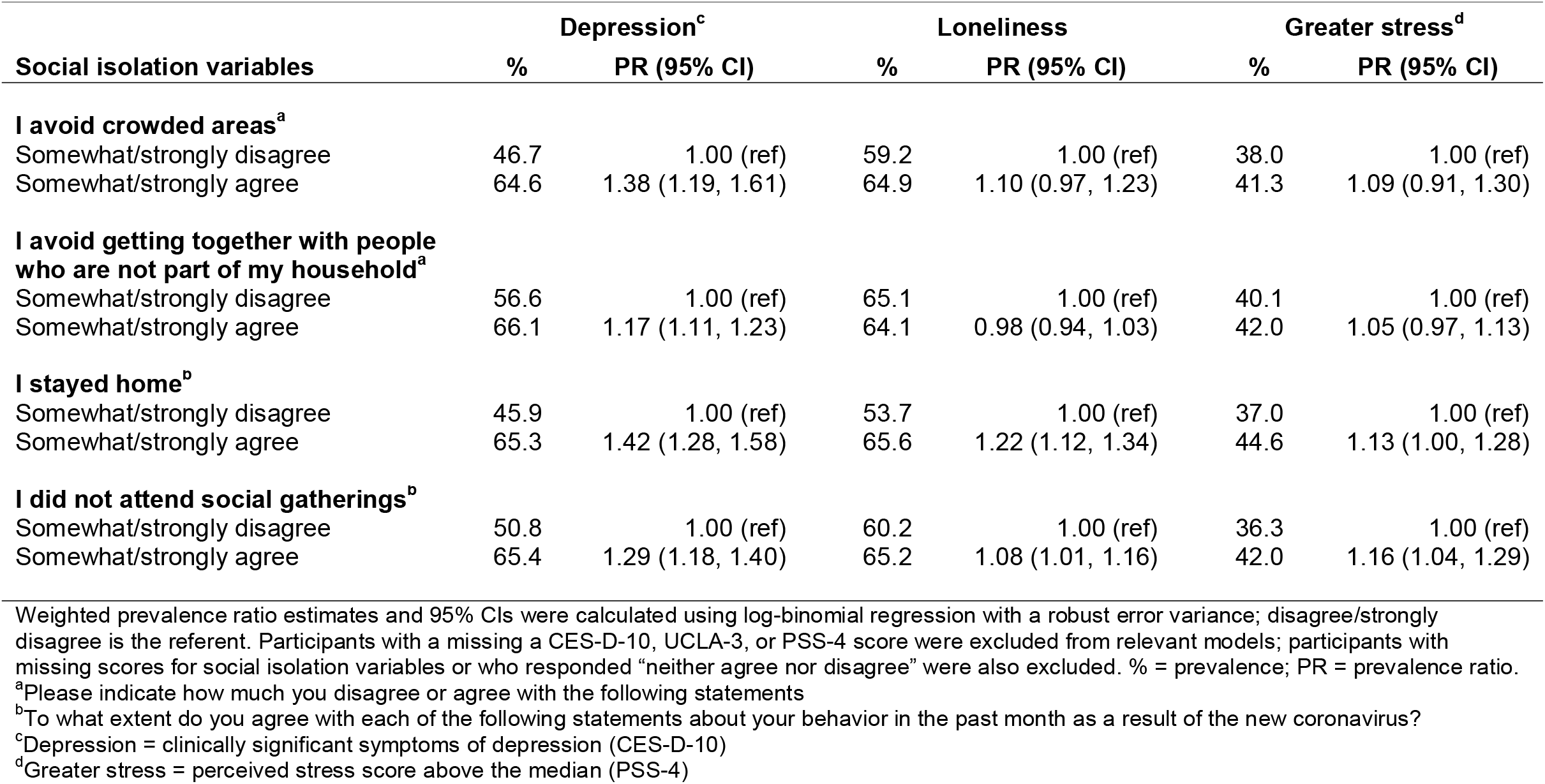
Associations between other social isolation variables and psychological distress outcomes

| Social isolation variables | Depression<sup>c</sup> |  | Loneliness |  | Greater stress<sup>d</sup> |  |
| --- | --- | --- | --- | --- | --- | --- |
|  | % | PR (95% CI) | % | PR (95% CI) | % | PR (95% CI) |
| **I avoid crowded areas<sup>a</sup>** |  |  |  |  |  |  |
| Somewhat/strongly disagree | 46.7 | 1.00 (ref) | 59.2 | 1.00 (ref) | 38.0 | 1.00 (ref) |
| Somewhat/strongly agree | 64.6 | 1.38 (1.19, 1.61) | 64.9 | 1.10 (0.97, 1.23) | 41.3 | 1.09 (0.91, 1.30) |
| **I avoid getting together with people who are not part of my household<sup>a</sup>** |  |  |  |  |  |  |
| Somewhat/strongly disagree | 56.6 | 1.00 (ref) | 65.1 | 1.00 (ref) | 40.1 | 1.00 (ref) |
| Somewhat/strongly agree | 66.1 | 1.17 (1.11, 1.23) | 64.1 | 0.98 (0.94, 1.03) | 42.0 | 1.05 (0.97, 1.13) |
| **I stayed home<sup>b</sup>** |  |  |  |  |  |  |
| Somewhat/strongly disagree | 45.9 | 1.00 (ref) | 53.7 | 1.00 (ref) | 37.0 | 1.00 (ref) |
| Somewhat/strongly agree | 65.3 | 1.42 (1.28, 1.58) | 65.6 | 1.22 (1.12, 1.34) | 44.6 | 1.13 (1.00, 1.28) |
| **I did not attend social gatherings<sup>b</sup>** |  |  |  |  |  |  |
| Somewhat/strongly disagree | 50.8 | 1.00 (ref) | 60.2 | 1.00 (ref) | 36.3 | 1.00 (ref) |
| Somewhat/strongly agree | 65.4 | 1.29 (1.18, 1.40) | 65.2 | 1.08 (1.01, 1.16) | 42.0 | 1.16 (1.04, 1.29) |

Weighted prevalence ratio estimates and 95% CIs were calculated using log-binomial regression with a robust error variance; disagree/strongly disagree is the referent. Participants with a missing a CES-D-10, UCLA-3, or PSS-4 score were excluded from relevant models; participants with missing scores for social isolation variables or who responded "neither agree nor disagree" were also excluded. % = prevalence; PR = prevalence ratio.
<sup>a</sup>Please indicate how much you disagree or agree with the following statements
<sup>b</sup>To what extent do you agree with each of the following statements about your behavior in the past month as a result of the new coronavirus?
<sup>c</sup>Depression = clinically significant symptoms of depression (CES-D-10)
<sup>d</sup>Greater stress = perceived stress score above the median (PSS-4)

**Figure 1.**
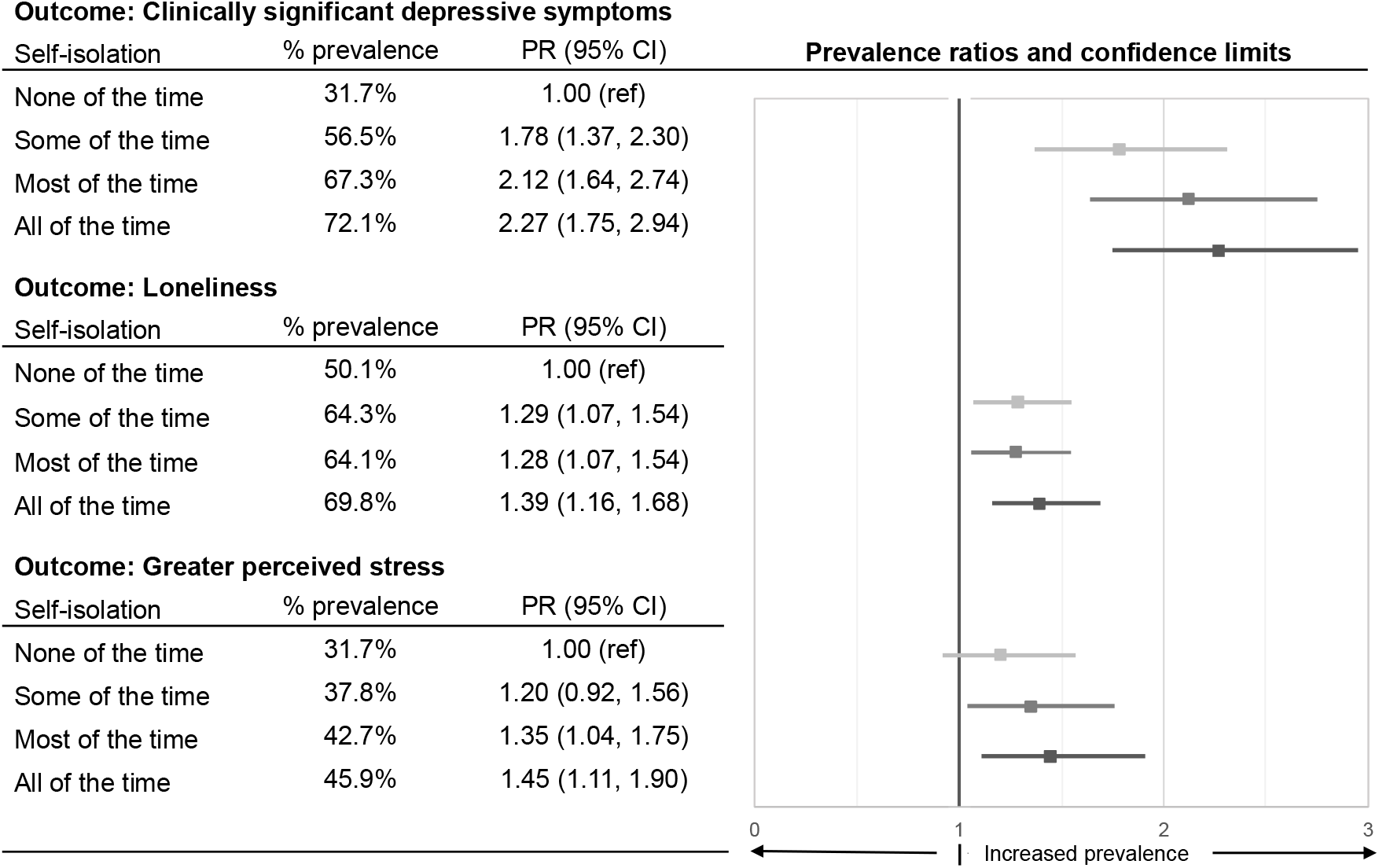
Associations between level of self-isolation and psychological distress outcomes Notes: PR=prevalence ratio; Weighted prevalence ratio estimates and 95% CIs were calculated using log-binomial regression with a robust error variance; self-isolation: none of the time is the referent; scores for the Perceived Stress Scale (PSS-4) were dichotomized at the median

Supplemental materials contain unweighted exposure and outcome distributions (Appendix A) and expanded tables containing weighted and unweighted estimates for associations between all social isolation variables and psychological distress outcomes that include the number of participants with a given outcome within each exposure category (Appendix B). Lastly, we provided weighted and unweighted estimates for associations between self-isolation and psychological distress outcomes stratified by age, race and ethnicity, gender, and student type (Appendix C).

## DISCUSSION

Prevalence of psychological distress outcomes among a cohort of undergraduate, graduate, and professional students in the southern United States in June 2020 was strikingly high. Clinically significant levels of symptoms of depression were reported by almost two-thirds (64%) of students. This compares to global data, which has shown significant increases in reports of depression among all age groups due to the pandemic (14,15). Although measures and criteria differ, in a study among adults in the United States, depressive symptoms were more than 3-fold higher during COVID-19 compared with before the pandemic (27.8% vs. 8.5%) (16). Similarly, in a nationally representative survey among US adults in June 2020, reports of appreciable symptoms of depressive disorder were approximately four times higher than in 2019 (24.3% vs. 6.5%) (1,17). Further, in June 2020, reports were higher among young people 18-24 years (52.3%) than any age group (1).

Among UNC-CH students, almost two-thirds (65%) were categorized as lonely. In a cross-cohort analysis of data from UK adults, 39% were categorized as lonely using the same UCLA-3 cut point during the pandemic compared to 26% before the pandemic (18). There is consistent evidence linking loneliness to poor physical and mental health outcomes, particularly among young people (19–21). For example, a rapid review on the impact of social isolation and loneliness on the mental health of children, adolescents, and young adults found that loneliness for long durations was associated with depression, anxiety, and posttraumatic stress (22). Reducing feelings of loneliness is also crucial to preventing suicide (23–25). Further, while our measure of perceived stress was dichotomized at the sample median, in a recent survey conducted among college students in Texas, 71% of students indicated that their stress levels had increased during the pandemic (26).

Our study found that level of self-reported self-isolation was associated with clinically significant depressive symptoms, loneliness, and greater perceived stress, with the largest effect estimates observed for depression. These findings are consistent with previous research that has demonstrated the profound impact of social isolation on mental health (19,20,27). In a survey conducted in Australia, adolescents’ greatest concerns during COVID-19 were around the disruption to their social interactions and activities, and that feeling socially disconnected was associated with increased anxiety and depression, and decreased life satisfaction (28). Therefore, for younger age groups, the impacts of social isolation on mental health may be particularly pronounced. In the era of COVID-19, social isolation is a widely shared experience despite high levels of social media use, particularly among young people. Data from our study and others suggest that confinement in place and lack of real-time, face-to-face human contact may themselves degrade our mental health.

Interventions to support students experiencing psychological distress during COVID-19 are critical given that physical distancing measures are expected to continue (6). Mental health disorders can negatively impact a student’s academic success (29), in addition to their general health and well-being. There is a growing body of evidence demonstrating the efficacy of peer support interventions for depression (30,31), with some evidence among university students (32). Peer support may have value among university students who show low engagement in traditional mental health services (2,33). Universities should also promote evidence-based initiatives aimed at reducing the psychological impact of COVID-19, including providing access to telepsychology services (34) and promoting personal strategies to improve one’s mental health (e.g., connecting with others, engaging in hobbies or physical activity, practicing meditation, etc.) (14,35). Last, universities should emphasize that students make efforts to safely connect with others while pandemic control measures are in place (e.g., meeting virtually or outdoors in small numbers while wearing masks).

There are several limitations of this research. First, this study used weighting to make inferences about the UNC-CH student population; findings, however, may not be generalizable to US college students more broadly. Second, we utilized clinically validated screening instruments to assess symptoms of mental health disorders and psychological distress; diagnostic evaluations were not conducted. Next, the survey administration coincided with protests in support of the Black Lives Matter movement across the United States. This may have impacted student responses and confounded our analysis given that Black/African American students reported a slightly higher prevalence of clinically significant depressive symptoms (67%). Lastly, although weighting methods were used to adjust for nonresponse, their effectiveness is limited if there are differences between survey respondents and non-respondents on study variables not accounted for by the weighting.

## CONCLUSIONS

The prevalence of adverse mental health outcomes among a cohort of college students in the southern United States was exceptionally high, with 64% of students reporting clinically significant depressive symptoms. Given that college and university students represent approximately 6% of the US population, the findings of this research document a significant burden of mental health distress. University policies to address this disparity in psychological distress should expand utilization of telepsychology services and ensure safe access to clinical treatment service options, as well as promote strategies for social connectedness and personal wellness. Research examining the long-term impacts of social isolation on mental health among college students and how universities can prepare support systems to mitigate mental health consequences as the pandemic evolves are urgently needed.

## Data Availability

All data produced in the present study are available upon reasonable request to the authors

## Acknowledgements

This research was supported by the University of North Carolina at Chapel Hill and grant F31MH119965 (PI: Giovenco) of the National Institutes of Mental Health. The funders had no role in study design, data collection and analysis, decision to publish, or preparation of the manuscript.

