## Appendices for "Social isolation and psychological distress among southern US college students in the era of COVID-19"

### APPENDIX A. Unweighted distributions of social isolation exposure variables and psychological distress outcome variables

**Table A.1. Unweighted distribution of social isolation exposure variables (N=7,012)**

| Social isolation variables | N (%) | 95% CI |
|---|---|---|
| **Self-isolation[a]** | | |
| None of the time | 132 (2%) | 1.6%, 2.2% |
| Some of the time | 2483 (35%) | 34.3%, 36.6% |
| Most of the time | 3507 (50%) | 48.9%, 51.2% |
| All of the time | 882 (13%) | 11.8%, 13.4% |
| **I avoid crowded areas[b]** | | |
| Somewhat/strongly disagree | 227 (3%) | 2.9%, 3.8% |
| Somewhat/strongly agree | 6519 (97%) | 96.2%, 97.1% |
| **I avoid getting together with people who are not part of my household[b]** | | |
| Somewhat/strongly disagree | 1395 (22%) | 21.1%, 23.2% |
| Somewhat/strongly agree | 4896 (78%) | 76.8%, 78.9% |
| **I stayed home[c]** | | |
| Somewhat/strongly disagree | 489 (7%) | 6.6%, 7.8% |
| Somewhat/strongly agree | 6322 (93%) | 92.2%, 93.4% |
| **I did not attend social gatherings[c]** | | |
| Somewhat/strongly disagree | 647 (10%) | 8.9%, 10.4% |
| Somewhat/strongly agree | 6053 (90%) | 89.6%, 91.1% |

Estimates exclude 8 (.11%) participants missing self-isolation; 21 (.30%) missing and 245 (3.5%) who responded “neither agree nor disagree” for “I avoid crowded areas”; 16 (.23%) missing and 705 (10.1%) who responded “neither agree nor disagree” for “I avoid getting together with people who are not in my household”; 14 (.20%) missing and 187 (2.7%) who responded “neither agree nor disagree” for “I stayed home”; and 25 (.36%) missing and 287 (4.1%) who responded “neither agree nor disagree” for “I did not attend social gatherings”

[a]To what extent are you self-isolating?

[b]Please indicate how much you disagree or agree with the following statements

[c]To what extent do you agree with each of the following statements about your behavior in the past month as a result of the new coronavirus?

**Table A.2. Unweighted distribution of psychological distress outcome variables (N=7,012)**

| Mental health variables | N (%) or Median (IQR) | Mean (95% CI) or 95% CI |
|---|---|---|
| **CES-D-10** | | |
| Non-clinically significant depressive symptoms (score <10) | 2378 (34%) | 33.4%, 35.6% |
| Clinically significant depressive symptoms (score ≥10) | 4520 (66%) | 64.4%, 66.6% |
| **UCLA-3** | | |
| Not lonely (score=3-5) | 2335 (33%) | 32.3%, 34.5% |
| Lonely (score=6-9) | 4653 (67%) | 65.5%, 67.7% |
| **PSS-4** | | |
| Lower stress (score=0-8) | 3922 (56%) | 55.1%, 57.5% |
| Greater stress (score=9-16) | 3046 (44%) | 42.5%, 44.9% |

Estimates exclude 114 (1.6%) participants missing a CES-D-10 score, 24 (.34%) missing a UCLA-3 score, and 44 (.63%) missing a PSS-4 score; scores for the Perceived Stress Scale (PSS-4) were dichotomized at the median

### APPENDIX B. Associations between social isolation variables and psychological distress outcomes, unweighted and weighted estimates and sample sizes

**Table B.1. Associations between social isolation variables and psychological distress outcomes, weighted counts, prevalence estimates, and prevalence ratios**

| | | Depression[d] | | | Loneliness | | | Greater stress[e] | | |
|---|---|---|---|---|---|---|---|---|---|---|
| **Social isolation variables** | **N (%)** | **N / Total N** | **%** | **PR (95% CI)** | **N / Total N** | **%** | **PR (95% CI)** | **N / Total N** | **%** | **PR (95% CI)** |
| **Self-isolation[a]** | | | | | | | | | | |
| None of the time | 659 (2%) | 209/659 | 31.7 | 1.00 (ref) | 330/659 | 50.1 | 1.00 (ref) | 206/650 | 31.7 | 1.00 (ref) |
| Some of the time | 11288 (34%) | 6292/11137 | 56.5 | 1.78 (1.37, 2.30) | 7239/11254 | 64.3 | 1.29 (1.07, 1.54) | 4245/11220 | 37.8 | 1.20 (0.92, 1.56) |
| Most of the time | 16927 (51%) | 11181/16617 | 67.3 | 2.12 (1.64, 2.74) | 10808/16860 | 64.1 | 1.28 (1.07, 1.54) | 7174/16802 | 42.7 | 1.35 (1.04, 1.75) |
| All of the time | 4324 (13%) | 3056/4236 | 72.1 | 2.27 (1.75, 2.94) | 3000/4301 | 69.8 | 1.39 (1.16, 1.68) | 1973/4295 | 45.9 | 1.45 (1.11, 1.90) |
| **I avoid crowded areas[b]** | | | | | | | | | | |
| Somewhat/strongly disagree | 1065 (3%) | 494/1057 | 46.7 | 1.00 (ref) | 628/1061 | 59.2 | 1.00 (ref) | 400/1053 | 38.0 | 1.00 (ref) |
| Somewhat/strongly agree | 30890 (97%) | 19631/30385 | 64.6 | 1.38 (1.19, 1.61) | 19972/30774 | 64.9 | 1.10 (0.97, 1.23) | 12684/30681 | 41.3 | 1.09 (0.91, 1.30) |
| **I avoid getting together with people who are not part of my household[b]** | | | | | | | | | | |
| Somewhat/strongly disagree | 6416 (21%) | 3576/6319 | 56.6 | 1.00 (ref) | 4165/6398 | 65.1 | 1.00 (ref) | 2555/6368 | 40.1 | 1.00 (ref) |
| Somewhat/strongly agree | 23459 (79%) | 15237/23049 | 66.1 | 1.17 (1.11, 1.23) | 14977/23376 | 64.1 | 0.98 (0.94, 1.03) | 9797/23300 | 42.0 | 1.05 (0.97, 1.13) |
| **I stayed home[c]** | | | | | | | | | | |
| Somewhat/strongly disagree | 2352 (7%) | 1062/2315 | 45.9 | 1.00 (ref) | 1254/2335 | 53.7 | 1.00 (ref) | 866/2338 | 37.0 | 1.00 (ref) |
| Somewhat/strongly agree | 29931 (93%) | 19227/29443 | 65.3 | 1.42 (1.28, 1.58) | 19576/29846 | 65.6 | 1.22 (1.12, 1.34) | 12464/27942 | 44.6 | 1.13 (1.00, 1.28) |
| **I did not attend social gatherings[c]** | | | | | | | | | | |
| Somewhat/strongly disagree | 3046 (10%) | 1517/2985 | 50.8 | 1.00 (ref) | 1830/3039 | 60.2 | 1.00 (ref) | 1095/3016 | 36.3 | 1.00 (ref) |
| Somewhat/strongly agree | 28724 (90%) | 18476/28266 | 65.4 | 1.29 (1.18, 1.40) | 18660/28632 | 65.2 | 1.08 (1.01, 1.16) | 11989/28561 | 42.0 | 1.16 (1.04, 1.29) |

Weighted prevalence ratio estimates and 95% CIs were calculated using log-binomial regression with a robust error variance; none of the time and disagree/strongly disagree are reference groups. Participants with a missing a CES-D-10, UCLA-3, or PSS-4 score were excluded from relevant models; participants with missing scores for social isolation variables or who responded "neither agree nor disagree" were also excluded. % = prevalence; PR = prevalence ratio.

[a]To what extent are you self-isolating?

[b]Please indicate how much you disagree or agree with the following statements

[c]To what extent do you agree with each of the following statements about your behavior in the past month as a result of the new coronavirus?

[d]Depression = clinically significant symptoms of depression (CES-D-10)

[e]Greater stress = perceived stress score above the median (PSS-4)

**Table B.2. Associations between social isolation variables and psychological distress outcomes, unweighted counts, prevalence estimates, and prevalence ratios**

| | | Depression[d] | | | Loneliness | | | Greater stress[e] | | |
|---|---|---|---|---|---|---|---|---|---|---|
| **Social isolation variables** | **N (%)** | **N / Total N** | **%** | **PR (95% CI)** | **N / Total N** | **%** | **PR (95% CI)** | **N / Total N** | **%** | **PR (95% CI)** |
| **Self-isolation[a]** | | | | | | | | | | |
| None of the time | 132 (2%) | 47/132 | 35.6 | 1.00 (ref) | 70/132 | 53.0 | 1.00 (ref) | 46/130 | 35.4 | 1.00 (ref) |
| Some of the time | 2483 (35%) | 1442/2447 | 58.9 | 1.66 (1.31, 2.09) | 1639/2477 | 66.2 | 1.25 (1.06, 1.47) | 1001/2470 | 40.5 | 1.15 (0.90, 1.45) |
| Most of the time | 3507 (50%) | 2394/3448 | 69.4 | 1.95 (1.55, 2.46) | 2314/3495 | 66.2 | 1.25 (1.06, 1.47) | 1571/3485 | 45.1 | 1.27 (1.01, 1.61) |
| All of the time | 882 (12%) | 633/865 | 73.2 | 2.06 (1.63, 2.59) | 625/878 | 71.2 | 1.34 (1.14, 1.58) | 426/877 | 48.6 | 1.37 (1.08, 1.75) |
| **I avoid crowded areas[b]** | | | | | | | | | | |
| Somewhat/strongly disagree | 227 (3%) | 117/225 | 52.0 | 1.00 (ref) | 138/226 | 61.1 | 1.00 (ref) | 96/244 | 39.3 | 1.00 (ref) |
| Somewhat/strongly agree | 6519 (97%) | 4267/6416 | 66.5 | 1.28 (1.13, 1.45) | 4346/6499 | 66.9 | 1.10 (0.99, 1.22) | 2836/6482 | 43.8 | 1.02 (0.88, 1.19) |
| **I avoid getting together with people who are not part of my household[b]** | | | | | | | | | | |
| Somewhat/strongly disagree | 1395 (22%) | 815/1375 | 59.3 | 1.00 (ref) | 927/1392 | 66.6 | 1.00 (ref) | 599/1386 | 43.2 | 1.00 (ref) |
| Somewhat/strongly agree | 4896 (78%) | 3277/4813 | 68.1 | 1.15 (1.10, 1.21) | 3229/4882 | 66.1 | 0.99 (0.95, 1.04) | 2157/4867 | 44.3 | 1.03 (0.96, 1.10) |
| **I stayed home[c]** | | | | | | | | | | |
| Somewhat/strongly disagree | 489 (7%) | 233/481 | 48.4 | 1.00 (ref) | 269/487 | 55.2 | 1.00 (ref) | 195/486 | 40.1 | 1.00 (ref) |
| Somewhat/strongly agree | 6322 (93%) | 4184/6223 | 67.2 | 1.39 (1.26, 1.52) | 4262/6305 | 67.6 | 1.22 (1.13, 1.33) | 2785/6288 | 44.3 | 1.10 (0.99, 1.23) |
| **I did not attend social gatherings[c]** | | | | | | | | | | |
| Somewhat/strongly disagree | 647 (10%) | 342/634 | 53.9 | 1.00 (ref) | 400/646 | 61.9 | 1.00 (ref) | 254/641 | 39.6 | 1.00 (ref) |
| Somewhat/strongly agree | 6053 (90%) | 4010/5960 | 67.3 | 1.25 (1.16, 1.34) | 4056/6036 | 67.2 | 1.09 (1.02, 1.16) | 2670/6024 | 44.3 | 1.12 (1.01, 1.24) |

Unweighted prevalence ratio estimates and 95% CIs were calculated using log-binomial regression; None of the time and disagree/strongly disagree are reference groups. Participants with a missing a CES-D-10, UCLA-3, or PSS-4 score were excluded from relevant models; participants with missing scores for social isolation variables or who responded “neither agree nor disagree” were also excluded. % = prevalence; PR = prevalence ratio.

[a]To what extent are you self-isolating?

[b]Please indicate how much you disagree or agree with the following statements

[c]To what extent do you agree with each of the following statements about your behavior in the past month as a result of the new coronavirus?

[d]Depression = clinically significant symptoms of depression (CES-D-10)

[e]Greater stress = perceived stress score above the median (PSS-4)

### APPENDIX C. Weighted and unweighted associations between self-isolation and psychological distress outcomes stratified by age, race/ethnicity, gender, and student type

**Table C.1. Associations between self-isolation and psychological distress outcomes stratified by sample characteristics, weighted counts, prevalence estimates, and prevalence ratios**

| Characteristic | Depression[a] | Self-isolating most or all of the time vs some or none of the time | Loneliness | Self-isolating most or all of the time vs some or none of the time | Greater stress[b] | Self-isolating most or all of the time vs some or none of the time |
|---|---|---|---|---|---|---|
| | N (%) | PR (95% CI) | N (%) | PR (95% CI) | N (%) | PR (95% CI) |
| Age | | | | | | |
| <21 years | 9517 (62%) | 1.23 (1.17, 1.31) | 11042 (71%) | 1.08 (1.03, 1.13) | 6564 (43%) | 1.15 (1.06, 1.25) |
| 21-24 years | 5623 (67%) | 1.22 (1.13, 1.33) | 5667 (67%) | 1.05 (0.97, 1.13) | 3779 (44%) | 1.21 (1.07, 1.37) |
| 25-34 years | 4401 (63%) | 1.27 (1.13, 1.42) | 3765 (54%) | 1.07 (0.95, 1.21) | 2630 (38%) | 1.14 (0.96, 1.35) |
| ≥35 years | 1216 (59%) | 1.36 (1.03, 1.80) | 929 (44%) | 0.91 (0.68, 1.23) | 637 (30%) | 1.74 (1.05, 2.87) |
| Race/Ethnicity | | | | | | |
| White | 11931 (63%) | 1.25 (1.19, 1.32) | 12527 (65%) | 1.03 (0.98, 1.08) | 7775 (41%) | 1.14 (1.06, 1.23) |
| Black or African American | 1733 (67%) | 1.15 (0.96, 1.38) | 1628 (62%) | 1.13 (0.93, 1.38) | 1103 (42%) | 1.28 (0.94, 1.73) |
| Asian | 3243 (62%) | 1.31 (1.12, 1.52) | 3289 (62%) | 1.06 (0.93, 1.20) | 2058 (39%) | 1.40 (1.11, 1.77) |
| Hispanic of any race | 1848 (65%) | 1.32 (1.12, 1.55) | 1859 (65%) | 1.07 (0.93, 1.24) | 1345 (47%) | 1.11 (0.90, 1.37) |
| Other or multiple races | 1134 (66%) | 1.06 (0.89, 1.25) | 1168 (68%) | 0.94 (0.80, 1.09) | 759 (44%) | 0.98 (0.76, 1.26) |
| Gender | | | | | | |
| Women | 13464 (71%) | 1.16 (1.12, 1.21) | 13048 (67%) | 1.00 (0.96, 1.05) | 9326 (48%) | 1.11 (1.05, 1.19) |
| Men | 7276 (54%) | 1.37 (1.24, 1.50) | 8340 (61%) | 1.06 (0.98,1.14) | 4265 (31%) | 1.22 (1.06, 1.40) |
| Student type | | | | | | |
| Undergraduate | 13265 (63%) | 1.24 (1.18, 1.30) | 14790 (70%) | 1.05 (1.01, 1.10) | 9179 (43%) | 1.14 (1.07, 1.23) |
| Graduate/prof | 7494 (64%) | 1.25 (1.15, 1.36) | 6613 (55%) | 1.04 (0.96, 1.14) | 4430 (37%) | 1.26 (1.10, 1.44) |

Weighted prevalence ratio estimates and 95% CIs were calculated using log-binomial regression with a robust error variance; self-isolation: some or none of the time is the referent. Participants with a missing a CES-D-10, UCLA-3, or PSS-4 score were excluded from relevant models; participants with missing age, race/ethnicity, gender, or a score for self-isolation were also excluded. PR = prevalence ratio.

[a]Depression = clinically significant symptoms of depression (CES-D-10)

[b]Greater stress = perceived stress score above the median (PSS-4)

**Table C.2. Associations between self-isolation and psychological distress outcomes stratified by sample characteristics, unweighted counts, prevalence estimates, and prevalence ratios**

| Characteristic | Depression[a] | Self-isolating most or all of the time vs some or none of the time | Loneliness | Self-isolating most or all of the time vs some or none of the time | Greater stress[b] | Self-isolating most or all of the time vs some or none of the time |
|---|---|---|---|---|---|---|
| | N (%) | PR (95% CI) | N (%) | PR (95% CI) | N (%) | PR (95% CI) |
| Age | | | | | | |
| <21 years | 2318 (64%) | 1.20 (1.14, 1.26) | 2637 (72%) | 1.07 (1.03, 1.12) | 1638 (45%) | 1.12 (1.04, 1.21) |
| 21-24 years | 1152 (68%) | 1.23 (1.14, 1.33) | 1158 (68%) | 1.04 (0.97, 1.12) | 797 (47%) | 1.22 (1.09, 1.36) |
| 25-34 years | 864 (66%) | 1.26 (1.14, 1.40) | 717 (54%) | 1.08 (0.96, 1.22) | 511 (39%) | 1.14 (0.97, 1.34) |
| ≥35 years | 185 (63%) | 1.31 (1.02, 1.67) | 140 (46%) | 0.91 (0.69, 1.19) | 100 (33%) | 1.66 (1.04, 2.65) |
| Race/Ethnicity | | | | | | |
| White | 2833 (65%) | 1.24 (1.18, 1.30) | 2954 (67%) | 1.03 (0.99, 1.08) | 1907 (43%) | 1.13 (1.06, 1.22) |
| Black or African American | 273 (69%) | 1.10 (0.94, 1.30) | 259 (64%) | 1.06 (0.89, 1.27) | 177 (44%) | 1.26 (0.95, 1.66) |
| Asian | 638 (64%) | 1.29 (1.12, 1.48) | 645 (64%) | 1.06 (0.94, 1.19) | 412 (41%) | 1.40 (1.13, 1.74) |
| Hispanic of any race | 356 (67%) | 1.22 (1.06, 1.40) | 359 (66%) | 1.06 (0.93, 1.21) | 261 (48%) | 1.05 (0.87, 1.27) |
| Other or multiple races | 248 (69%) | 1.02 (0.88, 1.18) | 252 (69%) | 0.93 (0.81, 1.06) | 173 (48%) | 0.97 (0.78, 1.21) |
| Gender | | | | | | |
| Women | 3462 (70%) | 1.17 (1.12, 1.21) | 3412 (68%) | 1.01 (0.97, 1.05) | 2425 (49%) | 1.11 (1.04, 1.18) |
| Men | 1053 (53%) | 1.36 (1.24, 1.50) | 1237 (62%) | 1.07 (0.99,1.15) | 617 (31%) | 1.21 (1.05, 1.39) |
| Student type | | | | | | |
| Undergraduate | 3061 (65%) | 1.21 (1.16, 1.26) | 3381 (71%) | 1.05 (1.01, 1.09) | 2181 (46%) | 1.13 (1.06, 1.21) |
| Graduate/prof | 1458 (66%) | 1.24 (1.15, 1.33) | 1271 (57%) | 1.04 (0.96, 1.13) | 865 (39%) | 1.25 (1.10, 1.41) |

Unweighted prevalence ratio estimates and 95% CIs were calculated using log-binomial regression; self-isolation: some or none of the time is the referent. Participants with a missing a CES-D-10, UCLA-3, or PSS-4 score were excluded from relevant models; participants with missing age, race/ethnicity, gender, or a score for self-isolation were also excluded. PR = prevalence ratio.

[a]Depression = clinically significant symptoms of depression (CES-D-10)

[b]Greater stress = perceived stress score above the median (PSS-4)
